# Accelerated hematopoietic aging in type 1 diabetes links telomere attrition to myocardial infarction

**DOI:** 10.64898/2026.05.13.26353138

**Authors:** Ziang Xu, Haibei Jiang, Tinsae Admassu, Skel Yeung, Sourav Roy, Campbell Ruffing, Michelle Shanguhyia, Zehra A. Syeda, David A. Alagpulinsa

## Abstract

Type 1 diabetes (T1D) is associated with early-onset and increased risk of coronary artery disease (CAD), particularly myocardial infarction (MI), that is not fully explained by traditional cardiovascular risk factors. Hematopoietic aging, marked by clonal hematopoiesis of indeterminate potential (CHIP) and leukocyte telomere length (LTL) attrition, has been linked to CAD risk in the general population. We investigated whether T1D is associated with increased prevalence and earlier onset of CHIP and accelerated LTL attrition, and whether these processes contribute to CAD risk.

We analyzed 416,565 UK Biobank participants (1,342 T1D cases) with harmonized CHIP calls and qPCR-derived LTL measurements. T1D was defined using ICD-10 codes, diagnosis before age 40 years, and insulin initiation within one year. CHIP was defined as variant allele fraction >=2%, with higher thresholds evaluated in sensitivity analyses. Multivariable regression compared CHIP prevalence, age-specific CHIP probability, and LTL between T1D and controls. Associations of CHIP and LTL with prevalent and incident (median follow-up approximately 13.5 years) MI and CAD were evaluated within T1D. Mendelian randomization, conjFDR, and integrated single-cell genomic analyses assessed genetic relationships.

CHIP prevalence was higher in T1D than in controls (4.40% vs 3.00%; absolute risk difference 1.39%, 95% CI 0.29 to 2.49; p=3.7×10-3), with higher adjusted odds (OR 1.71, 95% CI 1.31 to 2.23; p=6.9×10-5). Age modeling indicated an approximately 7-year earlier shift in CHIP risk in T1D (3% prevalence at age 52.2 vs 59.0 years). Mean LTL did not differ significantly between T1D and controls (beta −0.040 SD; p=0.13); however, T1D disease duration was associated with shorter LTL independent of covariates (beta −0.0082 SD per year; p=0.002). Within T1D, shorter LTL was associated with higher risk of both prevalent and incident MI (OR 0.77, 95% CI 0.65 to 0.91; p=0.003; HR 0.63, 95% CI 0.44 to 0.91; p=0.012). CHIP was not significantly associated with MI or CAD in this cohort. Genetic analyses supported a bidirectional relationship between T1D and LTL, but not CHIP, and identified 47 T1D-LTL loci enriched for hematopoietic stem and progenitor cell pathways.

In conclusion, T1D is associated with accelerated hematopoietic aging, reflected by earlier and more prevalent CHIP and disease duration-dependent LTL attrition. LTL attrition, but not CHIP, was associated with MI risk, implicating telomere dynamics as a contributor to excess cardiovascular risk in T1D.

## Introduction

Coronary artery disease (CAD) is the leading cause of morbidity and reduced lifespan among individuals with type 1 diabetes (T1D) [1–3]. In addition to increased risk, individuals with T1D experience CAD events at a younger age than the general population [1,4]. Particularly, individuals with T1D have 4- to 10-fold higher risk of myocardial infarction (MI) compared with those without diabetes even before 40 years of age [5–8]. Despite major advances in glycemic control and management of traditional cardiovascular risk factors, individuals with T1D continue to experience a substantial residual burden of CAD [9–13]. These observations suggest that there may be additional mechanisms beyond hyperglycemia and conventional risk factors that contribute to CAD in T1D [9,14–21].

Aging-related dysregulation of hematopoiesis has emerged as a novel independent modifier of CAD risk in the general population [22–24]. Specifically, clonal hematopoiesis of indeterminate potential (CHIP) and attrition of leukocyte telomere length (LTL) are complementary somatic hematopoietic aging processes that contribute to CAD risk independent of traditional cardiovascular risk factors [25–27]. CHIP arises from age-related acquisition of somatic mutations in hematopoietic stem and progenitor cells (HSPCs) that cause their clonal expansion and that of their progeny [28,29]. In parallel, LTL is a biological clock of hematopoiesis, reflecting the replicative history and cumulative proliferative stress of HSPCs [25,30–32]. CHIP and LTL are also bidirectionally connected, whereby longer LTL increases the risk of developing CHIP, while established CHIP drives LTL shortening [33,34].

Beyond CAD, prior studies have associated CHIP with risk of other age-related diseases, including autoimmune diseases such as rheumatoid arthritis [35] and metabolic diseases such as type 2 diabetes [36]. However, these adult-onset conditions typically develop over prolonged periods of subclinical immune activation, inflammation, and metabolic stress. These chronic perturbations impair HSPC integrity, promoting somatic clonal expansion and telomere attrition [31]. Consequently, CHIP detected shortly prior to disease diagnosis may reflect longstanding immune and metabolic stress rather than serving as a primary disease-initiating event.

An autoimmune and metabolic disease such as T1D, which is often diagnosed in children and young adults [37], provides an opportunity to evaluate the impact of chronic immune and metabolic stress across the life course on hematopoietic aging, and whether this contributes to CAD comorbidity. We previously showed that the genetic architecture of T1D overlaps with multiple hematopoietic traits, including leukocyte counts across multiple blood lineages, which were associated with risk of both T1D and CAD [38]. Independent studies have also shown that T1D risk variants preferentially localize to regulatory elements active across hematopoietic cell types, including HSPCs [39]. In T1D, the bone marrow microenvironment where HSPCs reside is also characterized by oxidative stress, DNA damage, chronic inflammation, and sympathetic denervation, along with glycemic excursions [40–43]. These processes adversely impact HSPC integrity and promote clonal expansion and telomere attrition [31]. These observations support a model in which the genetic architecture and T1D-induced stress can adversely impact HSPC integrity and drive somatic evolution and LTL shortening to accelerate hematopoietic aging. However, whether T1D is associated with accelerated hematopoietic aging, reflected by earlier onset or increased prevalence of CHIP and accelerated LTL attrition, and whether these processes contribute to CAD risk has not been systematically investigated.

In this study, we test the hypothesis that T1D is associated with accelerated hematopoietic aging, characterized by earlier onset and increased prevalence of CHIP and accelerated telomere attrition, and that these processes may contribute to CAD risk.

## Methods

### Study population

We analyzed participants from the UK Biobank prospective cohort [44] with available harmonized somatic CHIP calls and LTL measurements at baseline. Individual-level data, CHIP calls, and qPCR-derived LTL measurements were accessed through an approved UK Biobank application (project ID: 214427). The UK Biobank was approved by the Northwest Multi-centre Research Ethics Committee as a research tissue bank, and all participants provided written informed consent [45,46].

T1D was defined using International Classification of Diseases, Tenth Revision (ICD-10) diagnostic codes (E10.x), diagnosis before 40 years of age, and insulin initiation within one year of diagnosis.

Participants without diabetes served as controls. Individuals with diabetes diagnoses not meeting the T1D definition, as well as those with prevalent hematologic malignancies or myeloid neoplasms at baseline, were excluded. Specifically, exclusions included ICD-10 diabetes diagnoses (E10–E14) not meeting the T1D case definition, hematologic malignancies or related neoplasms (C81–C96), myeloproliferative neoplasms (D45), myelodysplastic syndromes (D46), and neoplasms of uncertain or unknown behavior (D47).

### Definition of CHIP and LTL

CHIP, LTL, and covariates were assessed at the baseline blood draw; therefore, primary analyses represent cross-sectional associations at enrollment. CHIP was primarily defined as the presence of ≥1 somatic mutation in pre-specified CHIP-driver genes with variant allele fraction (VAF) ≥2%, based on UK Biobank–provided harmonized somatic mutation calls derived from blood DNA sequencing. Sensitivity analyses evaluated alternative thresholds (VAF ≥5%, ≥10%, and ≥20%) and categorical VAF groupings to assess dose–response relationships.

LTL was measured in UK Biobank using quantitative polymerase chain reaction (qPCR) and expressed as a standardized z-score. Lower z-scores indicate shorter telomeres and greater hematopoietic replicative aging.

### Cardiovascular outcomes

Myocardial infarction (MI) was the primary cardiovascular outcome, with coronary artery disease (CAD) as the secondary outcome. Outcomes were defined using curated UK Biobank phenotype algorithms supplemented by ICD-10 diagnostic and procedural codes. For incident analyses, follow-up accrued from the baseline assessment date until the first cardiovascular event, death, loss to follow-up, or administrative censoring. Participants with prevalent disease at baseline were excluded from incident models.

### Covariates

Multivariable models were adjusted for baseline demographic and genetic ancestry covariates, including age, age², sex, assessment center, and genetic principal components 1–10. Age was modeled using quadratic terms or restricted cubic splines to account for nonlinearity. Additional models further adjusted for smoking status and body mass index (BMI) in sensitivity analyses. Models including smoking and BMI were restricted to participants with complete data for these variables, resulting in a reduced analytic sample.

### Statistical analysis

We first evaluated whether T1D was associated with hematopoietic aging markers by comparing CHIP prevalence and LTL between individuals with T1D and diabetes-free controls. CHIP prevalence was analyzed using multivariable logistic regression models, and LTL was analyzed using multivariable linear regression models. Primary models were adjusted for age (continuous), sex, assessment center, and genetic ancestry (principal components 1–10), with additional models further adjusting for smoking status and body mass index (BMI) as sensitivity analyses.

To assess whether hematopoietic aging manifested earlier in individuals with T1D, we examined CHIP prevalence across predefined age strata and modeled predicted CHIP probability as a function of age using regression-based approaches. Age-dependent differences in LTL between T1D and control groups were evaluated using similarly specified regression models, and formal interaction terms were used to test whether associations between age and LTL differed by T1D status.

Within the T1D cohort, we evaluated whether cumulative disease exposure was associated with hematopoietic aging by testing associations of T1D duration with LTL and CHIP. Associations between disease duration and LTL were assessed using multivariable linear regression, whereas associations with CHIP prevalence were evaluated using multivariable logistic regression. Secondary analyses evaluated age at T1D diagnosis in relation to CHIP prevalence. We additionally examined associations between CHIP carrier status and LTL, including formal CHIP×T1D interaction testing to assess whether this relationship differed by T1D status.

Within the T1D cohort, we next evaluated associations between hematopoietic aging markers (CHIP and LTL) and cardiovascular outcomes. Prevalent MI and CAD were analyzed using multivariable logistic regression models. For incident analyses, participants with prevalent cardiovascular disease at baseline were excluded. Time-to-event analyses were performed using Cox proportional hazards models, with follow-up time defined from the baseline assessment visit to the first occurrence of the outcome of interest, death, loss to follow-up, or administrative censoring. The median follow-up time was 13.62 years (interquartile range [IQR] 10.58–14.70) for incident MI and 13.34 years (IQR 8.78–14.59) for incident CAD. The proportional hazards assumption was assessed using standard diagnostic methods.

All outcome models were adjusted for the same baseline covariates as described above. We first evaluated associations between CHIP and cardiovascular outcomes, followed by associations between LTL and cardiovascular outcomes. We then constructed joint models including both CHIP and LTL to assess their independent contributions to cardiovascular risk.

Sensitivity and secondary analyses included evaluating alternative CHIP variant allele fraction (VAF) thresholds (≥5%, ≥10%, and ≥20%), restricting analyses to participants free of cardiovascular disease at baseline, and additional adjustment for glycemic control (HbA1c), where available, as an exploratory sensitivity analysis. All statistical tests were two-sided, with statistical significance defined as P<0.05.

### Genome-wide genetic correlation analysis by cross-trait LDSC

We applied cross-trait linkage disequilibrium score regression (LDSC) to estimate genome-wide genetic correlations between T1D and hematopoietic aging traits [47], including LTL and CHIP. LDSC uses GWAS summary statistics to estimate shared genetic architecture attributable to common SNP effects across the genome.

Narrow-sense SNP heritability and pairwise genetic correlations were estimated from publicly available GWAS summary statistics for T1D (18,942 T1D cases and 501,638 controls) [48], LTL (472,174 healthy individuals) [49] and CHIP (25,657 cases and 342,869 controls) [50]. Analyses were performed using LDSC v1.0.1 with pre-computed LD scores derived from European populations of the 1000 Genomes Project Phase 3 reference panel. Variants were restricted to autosomal SNPs with minor allele frequency (MAF) >0.01. The extended major histocompatibility complex (MHC) region (chr6:25–34 Mb) was excluded due to its complex long-range LD structure.

Statistical significance was evaluated using two-sided tests, with P<0.05 considered nominally significant. Genetic correlation estimates were interpreted in the context of complementary cross-trait analyses, including conjunction FDR and Mendelian randomization.

### Conjunctional false discovery rate analysis

We applied conjunctional false discovery rate (*conjFDR*) analysis to identify pleiotropic loci influencing T1D and hematopoietic aging traits (CHIP and LTL). *ConjFDR* leverages cross-trait GWAS summary statistics to increase power for detecting pleiotropic variants while accounting for polygenic overlap between traits [51,52]. It identifies variants jointly associated with both traits regardless of effect direction, making it robust to opposing allelic effects within loci.

Conditional quantile–quantile (Q–Q) plots were first generated to evaluate enrichment of T1D-associated variants as a function of their significance in LTL or CHIP (and vice versa). Leftward deflection in conditional Q–Q plots was interpreted as evidence of polygenic overlap between traits. Conditional false discovery rate (*condFDR*) values were then calculated for each single nucleotide polymorphism (SNP), representing the posterior probability that a SNP is null for association with one trait conditional on its association with the other trait.

The *conjFDR* statistic was defined as the maximum of the two *condFDR* values for each SNP, providing a conservative estimate of the probability that a variant is null for either or both traits. SNPs with conjFDR < 0.05 were considered jointly associated with T1D and the corresponding hematopoietic aging trait. Lead variants were defined as the SNP with the lowest conjFDR value within each linkage disequilibrium (LD)-independent locus (r² < 0.1 within a ±250 kb window), with LD estimated using European reference data from the 1000 Genomes Project Phase 3.

To minimize spurious associations, GWAS summary statistics were pre-corrected for relevant covariates and population stratification. Analyses were restricted to autosomal SNPs, and a single representative signal was retained within the extended major histocompatibility complex (MHC) region (chr6:26–34 Mb, hg19) due to its complex LD structure. All analyses were performed using the pleioFDR R package [51,53] following established workflows.

Lead SNPs identified from conjFDR analyses were assigned to genes for annotation purposes based on genomic proximity, with each variant mapped to the nearest protein-coding gene using GRCh37/hg19 genomic coordinates [54]. Functional annotation of the mapped genes was subsequently performed through manual curation using publicly available databases, including Gene Ontology (GO), UniProt, and NCBI Gene, together with evidence from the published literature [55]. Genes were manually grouped into biologically relevant categories, including hematopoietic stem and progenitor cell regulation, immune signaling pathways, cell cycle and genomic stability, RNA regulation, and metabolic signaling. Where appropriate, gene functions were interpreted in the context of hematopoietic biology to facilitate biological interpretation of shared genetic loci.

### Integration of conjFDR loci with chromatin accessibility in HSPCs using SCAVENGE

To define the hematopoietic cellular context of shared genetic signals between T1D and LTL, we integrated conjFDR loci with single-cell chromatin accessibility (scATAC-seq) data from healthy donor bone marrow, obtained from a previously published dataset that encompasses all major HSPC populations, including hematopoietic stem cells (HSCs), multipotent progenitors (MPP/CMP-LMPP), common lymphoid progenitors (CLPs), granulocyte–monocyte progenitors (GMPs), and erythroid progenitors [56].

Trait–cell type localization was quantified using SCAVENGE (Single Cell Analysis of Variant Enrichment through Network propagation of Genomic data), a computational framework that maps trait-associated variants to their cellular context at single-cell resolution through network propagation [57]. SCAVENGE assigns per-cell trait relevance scores (TRS) by propagating genetic signal across a cell–cell similarity graph constructed from chromatin accessibility profiles. Lead variants representing T1D–LTL conjFDR loci (conjFDR < 0.05) were mapped to accessible chromatin regions by genomic overlap, and their regulatory potential was evaluated across single cells. The propagated signal was used to assign TRS to each cell, reflecting the relative extent to which its chromatin accessibility profile aligns with shared trait-associated regulatory elements.

TRS distributions were summarized across annotated HSPC populations to characterize preferential localization of shared T1D–LTL genetic signals across hematopoietic lineages. Visualization included projection of TRS onto a two-dimensional UMAP embedding of HSPCs and quantitative comparison of TRS distributions across cell populations.

To further resolve locus-level regulatory architecture, representative shared lead variants were examined in the context of aggregated chromatin accessibility profiles across HSPC populations. SNP-to-peak relationships were assessed based on genomic proximity and overlap with accessible regions to infer putative cis-regulatory mechanisms. Additional nearby variants displayed in locus plots represent surrounding GWAS signals for visualization purposes and were not used in SCAVENGE-based enrichment analyses.

### Mendelian randomization analyses

We performed bidirectional two-sample Mendelian randomization (MR) to evaluate potential causal relationships between T1D and between T1D and the hematopoietic aging traits LTL and CHIP. MR uses genetic variants robustly associated with an exposure as instrumental variables to estimate the causal effect of the exposure on an outcome.

Independent SNPs associated with each exposure at genome-wide significance (P < 5 × 10⁻⁸) were selected as instrumental variables and pruned using linkage disequilibrium (LD) clumping (r² < 0.001). Instrument strength was assessed using the F-statistic, with all instruments exceeding F>10. Exposure and outcome summary statistics were harmonized by aligning effect alleles across datasets, and ambiguous palindromic variants were excluded.

Primary causal estimates were obtained using the inverse-variance weighted (IVW) method under a multiplicative random-effects model, with SNP-specific effects calculated using the Wald ratio and combined across variants. Sensitivity analyses were performed using the weighted median and MR-Egger methods to evaluate robustness to potential horizontal pleiotropy. Cochran’s Q statistics were used to assess heterogeneity across instrumental variable estimates. The MR-Egger intercept test was used to assess evidence of directional pleiotropy. All MR analyses were conducted in R using the *TwoSampleMR package*.

## Results

### T1D is associated with increased prevalence and earlier onset of CHIP

We first evaluated whether individuals with T1D exhibit an increased burden of CHIP compared with diabetes-free controls in the UK Biobank. The analytic cohort comprised 1,342 individuals with rigorously defined T1D and 415,223 controls after exclusion of participants with any diabetes diagnosis or hematologic malignancy.

Across variant allele fraction (VAF) thresholds, CHIP prevalence was consistently higher in T1D, with the strongest differences observed at the conventional threshold used to define CHIP (VAF ≥2%). At this threshold, CHIP prevalence was 4.40% in T1D versus 3.00% in controls (absolute risk difference [ARD] +1.39%, 95% CI 0.29–2.49; p=3.7×10⁻³; **Figure 1a,b**; **Supplementary Table 1**). A similar enrichment was observed at VAF ≥5% (4.25% vs 2.91%; ARD +1.34%, 95% CI 0.26–2.42; p=4.6×10⁻³), whereas differences at higher clone sizes were directionally consistent but not statistically significant (VAF ≥10%: ARD +0.72%, p=0.071; VAF ≥20%: ARD +0.17%, p=0.589; **Figure 1b; Supplementary Table 1**), likely reflecting limited event counts and reduced statistical power among individuals with T1D. The prevalence of CHIP among control participants was consistent with prior estimates of ∼ 3–4% at VAF ≥2% [58,59].

**Figure 1.**
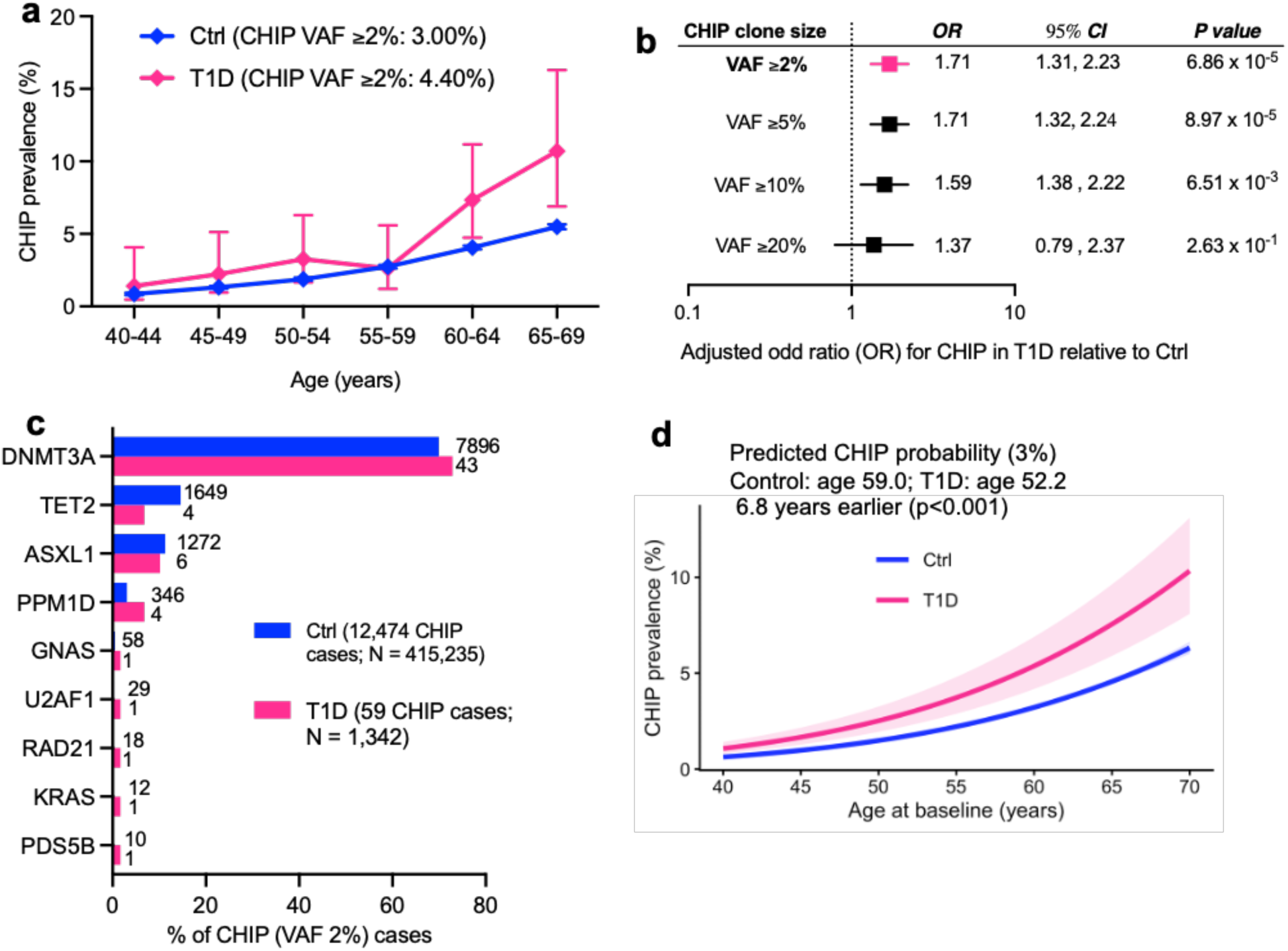
Increased prevalence and earlier onset of CHIP in T1D. (**a**) Age-stratified prevalence of clonal hematopoiesis of indeterminate potential (CHIP; variant allele fraction [VAF] ≥2%) in individuals with T1D and controls. Error bars indicate 95% confidence intervals. (**b**) Adjusted odds ratios (ORs) for CHIP in T1D versus controls across increasing clone size thresholds (VAF ≥2%, ≥5%, ≥10%, and ≥20%). (**c**) Distribution of CHIP driver genes among individuals with CHIP (VAF ≥2%) in T1D and controls, shown as the percentage of CHIP cases attributable to each gene. (**d**) Model-based predicted probability of CHIP by age, demonstrating earlier onset in T1D. The age at which 3% CHIP prevalence is reached occurs 6.8 years earlier in T1D compared with controls (52.2 vs 59.0 years; p<0.001). Shaded areas represent 95% confidence intervals.

In multivariable logistic regression adjusting for age, age², sex, ancestry principal components, smoking status, and body mass index (BMI), T1D remained associated with higher odds of CHIP at the conventional VAF threshold of ≥2% (OR 1.71, 95% CI 1.31–2.23; p=6.9×10⁻⁵; **Figure 1b**). Similar effect estimates were observed across increasing clone size thresholds, with statistically significant associations at VAF ≥5% and ≥10%, but not at VAF ≥20%, likely reflecting limited numbers of individuals harboring larger clones and consequent reduced statistical power. The effect estimates were highly stable across nested models, with minimal attenuation after adjustment for smoking or BMI individually or jointly (**Supplementary Table 2**).

The distribution of CHIP driver genes in individuals with T1D was broadly consistent with that observed in the general population, with DNMT3A mutations accounting for the majority of cases, followed by ASXL1 and TET2 (**Figure 1c; Supplementary Table 3**). The increased prevalence of CHIP in T1D was primarily driven by enrichment of DNMT3A-mutant clones (3.20% in T1D vs 1.90% in controls), whereas frequencies of other canonical CHIP drivers were similar between groups.

Because CHIP is strongly age-dependent, we examined whether T1D modifies the age trajectory of CHIP. Although individuals with T1D were modestly younger at recruitment (mean age 54.2±8.2 vs 56.2±8.1 years; **Supplementary Table 4**), age-modeled analyses demonstrated a clear upward shift in CHIP risk across the age spectrum (**Figure 1d**). Model-based predictions showed that individuals with T1D reached equivalent CHIP risk levels substantially earlier than controls (p<0.001). At a predicted CHIP probability of 3%, T1D participants reached this threshold at age 52.2 years compared with 59.0 years in controls (≈6.8 years earlier). At the 5% risk threshold, the corresponding ages were 59.0 versus 66.4 years, representing an approximately 7.4-year leftward shift in the age–CHIP curve. Consistent with a parallel upward displacement rather than differential age-dependent slopes, the T1D×age interaction was not significant (p=0.47). Among individuals who harbored CHIP, baseline age did not differ significantly between T1D and control groups after adjustment for sex and ancestry, indicating that the primary difference reflects earlier population-level emergence of CHIP rather than differences in age among CHIP carriers.

These findings demonstrate that T1D is associated with increased prevalence of CHIP and an approximately seven-year earlier shift in age-related CHIP risk, independent of major demographic, behavioral, and ancestry-related confounders.

### T1D disease duration is associated with shorter LTL but not CHIP

We analyzed LTL using the standardized UK Biobank phenotype (LTL_z) derived from quantitative PCR measurements and quality controlled by UK Biobank [60].

In unadjusted analyses, mean LTL_z was slightly lower in individuals with T1D compared with controls (0.004 vs 0.019), although overall distributions were highly similar between groups (**Figure 2a,b; Supplementary Table 5**). Median values were likewise comparable, indicating no substantial population-level shift in LTL associated with T1D.

**Figure 2.**
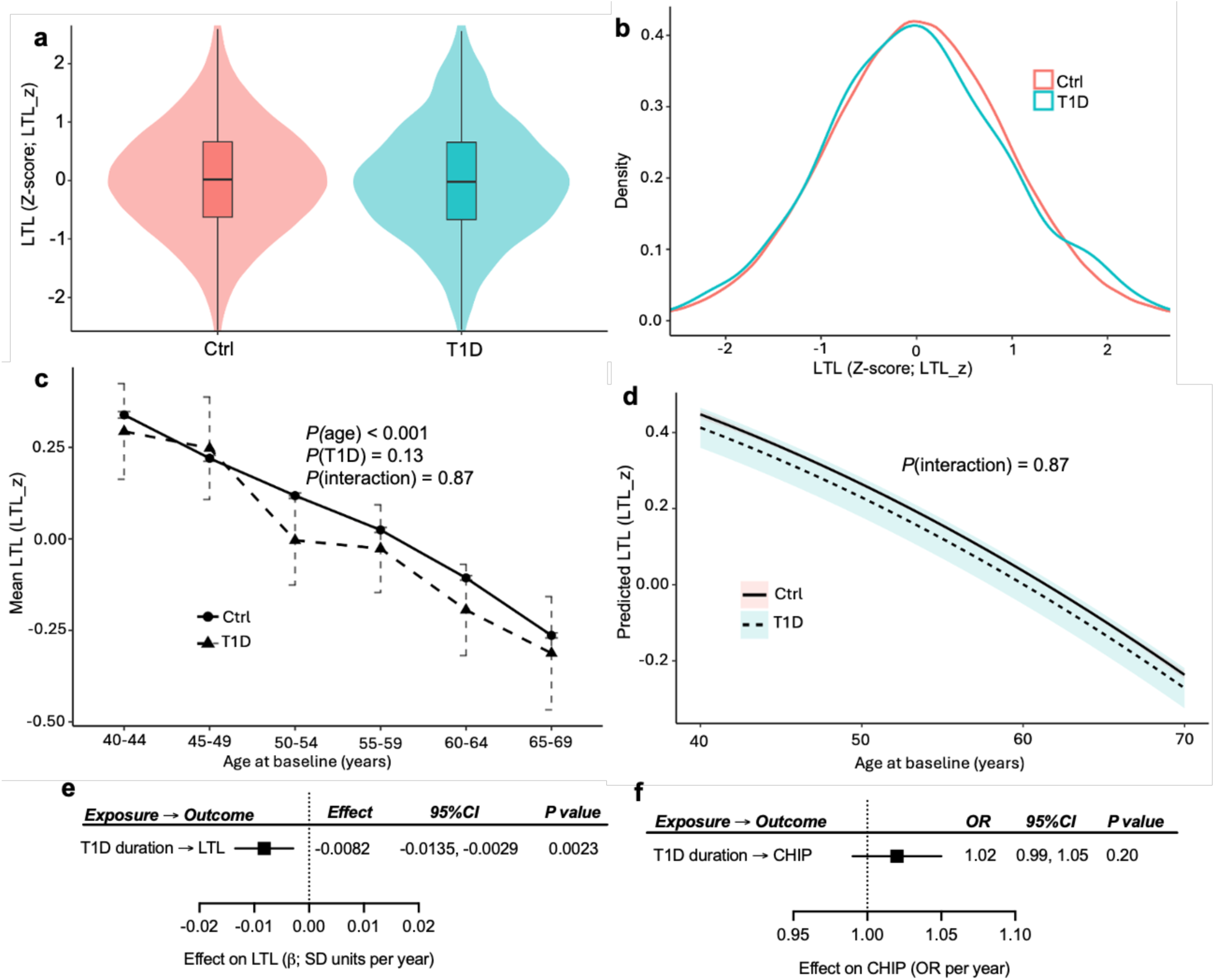
Leukocyte telomere length in type 1 diabetes. **(A)** Distribution of leukocyte telomere length (LTL). Violin plots of standardized LTL (LTL_Z) in individuals with type 1 diabetes (T1D) and controls. Central lines indicate medians and interquartile ranges. The overall distribution of LTL is similar between groups. **(B)** Density distribution of LTL by cohort. Kernel density plots of standardized LTL (LTL_Z) in T1D and control individuals demonstrate largely overlapping distributions. **(C)** Age-stratified LTL. Mean standardized LTL across age groups in T1D and control individuals. LTL declines with age in both groups, with broadly similar trajectories. **(D)** Model-based predicted LTL by age. Predicted standardized LTL as a function of age in T1D and control individuals from multivariable regression models adjusted for age, age², sex, genetic principal components, smoking, and body mass index. LTL declines with age in both groups, with no significant interaction between T1D status and age. **(e)** Association of T1D duration with hematopoietic aging measures. Longer T1D duration was associated with shorter leukocyte telomere length (LTL) (β = −0.0082 per year; p = 0.0023), indicating progressive telomere attrition. In contrast, T1D duration was not associated with clonal hematopoiesis of indeterminate potential (CHIP) prevalence (p = 0.20).

We performed multivariable linear regression to evaluate whether T1D was independently associated with LTL after accounting for demographic, behavioral, and ancestry factors. In models adjusting for age, age², sex, and ancestry principal components, T1D was associated with a small, non-significant reduction in LTL_z (β −0.040, 95% CI −0.092 to 0.011; p=0.127; **Supplementary Table 6**). The estimate remained stable after additional adjustment for smoking (β −0.039, p=0.136), BMI (β −0.036, p=0.176), and both jointly (β −0.035, p=0.188), with confidence intervals consistently crossing the null.

Given the strong age dependence of telomere length, we examined whether the relationship between T1D and LTL varied across the age spectrum. LTL significantly declined progressively with age in both groups (*P*(age) < 0.001), with no evidence of effect modification by T1D status (T1D×age interaction p=0.87; **Figure 2c**,**d**), indicating that age-related telomere attrition occurs in parallel in individuals with and without T1D.

Both CHIP and LTL are age-related but lifelong influences or exposures can shape their course. Therefore, we examined the impact of T1D disease duration on LTL and CHIP. Among individuals with T1D, longer disease duration was significantly associated with shorter LTL (**Figure 2e**). In multivariable models adjusting for age, age², sex, smoking, BMI, and ancestry principal components, each additional year of T1D disease duration was associated with a reduction of 0.0082 SD in LTL (β −0.0082 SD per year, 95% CI −0.0135 to −0.0029; p=0.0023; **Figure 2e; Supplementary Table 7**), consistent with disease-accelerated LTL attrition. In contrast, there was no evidence that T1D disease duration was associated with CHIP prevalence. In logistic regression models, longer disease duration was not significantly associated with CHIP (OR 1.02 per year, 95% CI 0.99–1.05; p=0.20; **Figure 2f; Supplementary Table 7**). Similarly, age at T1D diagnosis was not associated with CHIP risk (OR 0.98 per year, 95% CI 0.96–1.01; p=0.198; **Supplementary Table 7**). Although CHIP prevalence increased across duration categories descriptively, these differences were not statistically significant.

Because CHIP and LTL have been reported to exhibit bidirectional relationships, we examined whether CHIP carrier status was associated with shorter LTL and whether this relationship differed by T1D status. In the overall cohort, CHIP carriers had significantly shorter LTL than non-carriers (β −0.060 SD, 95% CI −0.077 to −0.042; p=1.7×10⁻¹¹; **Supplementary Table 8**). This association was essentially identical among controls but was not detectable within the T1D subgroup (β −0.033 SD, 95% CI −0.30 to 0.23; p=0.81). However, formal interaction testing provided no evidence that the CHIP–LTL relationship differed by T1D status (CHIP×T1D β +0.012 SD, 95% CI −0.24 to 0.27; p=0.92; **Supplementary Table 8**). Results were unchanged when age was modeled using restricted splines. These findings indicate that CHIP is robustly associated with shorter LTL in the general population, whereas within T1D this relationship was not detectable, likely reflecting limited statistical power and imprecise effect estimation due to the small number of T1D individuals harboring CHIP.

In summary, these results indicate that T1D exposure is associated with a significant acceleration of telomere attrition, whereas associations with CHIP burden were less evident, suggesting that LTL may represent a more sensitive marker of T1D-induced hematopoietic aging.

### Shorter LTL is associated with both prevalent and incident MI in T1D

We evaluated whether CHIP and LTL are associated with prevalent and incident MI and CAD among individuals with T1D. As an initial reference point, individuals with T1D exhibited a markedly increased baseline burden of MI compared with controls, irrespective of CHIP status (**Figure 3a**). The odds of prevalent MI were similarly elevated among individuals with T1D without CHIP (OR 3.60, 95% CI 3.02–4.28; p=1.53×10⁻⁴) and those with CHIP (OR 3.65, 95% CI 1.86–7.15; p=1.53×10⁻⁴), indicating that the excess MI risk in T1D is not explained by the presence of CHIP.

**Figure 3.**
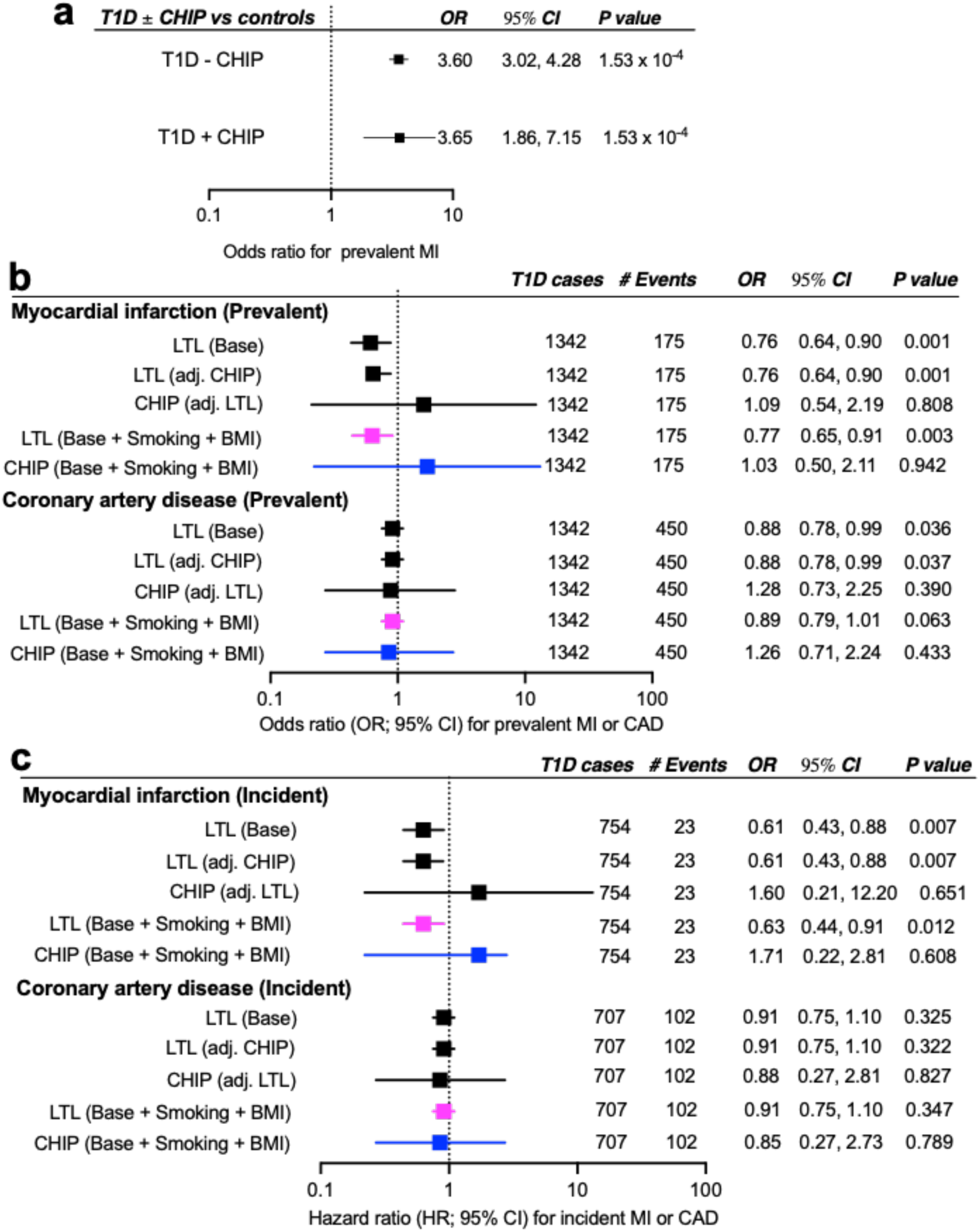
Shorter LTL, but not CHIP, is associated with MI risk in T1D. (**a**) Odds ratios (ORs) for prevalent myocardial infarction (MI) comparing individuals with type 1 diabetes (T1D) with and without clonal hematopoiesis of indeterminate potential (CHIP) to controls. (**b**) Associations of leukocyte telomere length (LTL) and CHIP with prevalent MI and coronary artery disease (CAD) within individuals with T1D. Models are shown with baseline adjustment and additional adjustment for smoking and body mass index (BMI). (**c**) Associations of LTL and CHIP with incident MI and CAD within individuals with T1D using Cox proportional hazards models. Hazard ratios (HRs) are shown with 95% confidence intervals (CIs) for baseline and fully adjusted models.

CHIP carrier status (VAF ≥2%) was not associated with prevalent MI or CAD across all models (**Figure 3b**). In contrast, LTL demonstrated a consistent inverse association with MI risk. Each 1-SD increase in LTL_z was associated with lower odds of prevalent MI (OR 0.76–0.77 across models; p=0.001–0.003; **Figure 3b**), with similar effect sizes after adjustment for CHIP, smoking, and BMI. For CAD, longer LTL was associated with lower odds in base models (OR 0.88, 95% CI 0.78–0.99; p=0.036) and after adjustment for CHIP (p=0.037). However, this association was attenuated and no longer statistically significant after further adjustment for smoking and BMI (OR 0.89, 95% CI 0.79–1.01; p=0.063). Across all models, CHIP remained unassociated with CAD. In joint models including both exposures, the association between LTL and MI remained unchanged, whereas CHIP remained null, indicating independent effects.

To assess longitudinal risk, we performed Cox proportional hazards analyses for incident cardiovascular events within the T1D cohort. Shorter LTL was associated with increased risk of incident MI (HR 0.61–0.63 per 1-SD longer LTL; p=0.007–0.012 across models; **Figure 3c**), whereas CHIP carriage was not associated with incident MI (HR 1.60–1.71; p>0.60). The association between LTL and incident MI remained consistent after adjustment for CHIP, smoking, and BMI. Neither LTL nor CHIP was significantly associated with incident CAD across models.

Taken together, these findings demonstrate that shorter LTL is associated with both prevalent and incident MI in T1D, whereas associations with CHIP were less evident. While LTL also showed a directionally consistent association with CAD, this relationship was attenuated after accounting for smoking and BMI. The stronger and more consistent relationship between LTL and MI risk suggests that LTL may represent a more sensitive and clinically relevant indicator of hematopoietic aging in T1D.

### T1D exhibits pervasive genetic relationships with LTL that converge on hematopoietic regulatory programs

To characterize the genetic architecture linking T1D with hematopoietic aging, we performed complementary genome-wide analyses, including cross-trait linkage disequilibrium score regression (LDSC), conjunctional false discovery rate (conjFDR), and bidirectional Mendelian randomization.

Cross-trait LDSC demonstrated a modest inverse genome-wide genetic correlation between T1D and LTL that did not reach significance under the unconstrained model (rg = −0.033, SE = 0.030, p = 0.26; **Supplementary Data 1**). Constraining the intercept to unity yielded a similar but nominally significant estimate (rg = −0.042, SE = 0.019, p = 0.026), indicating a weak global correlation sensitive to model assumptions. In contrast, no significant genome-wide genetic correlation was observed between T1D and CHIP, with point estimates directionally negative (rg = −0.018, SE = 0.052, p = 0.728; **Supplementary Data 1**).

Despite limited genome-wide correlation, conjFDR analysis revealed substantial localized genetic overlap between T1D and LTL, identifying 47 LD-independent loci jointly associated with both traits (conjFDR < 0.05; **Figure 4a; Supplementary Data 2**). These loci mapped to genes involved in HSPC maintenance and trafficking (e.g., *CXCR4, KIT, CD164, HHEX, FLI1*), immune regulation and lymphocyte differentiation (e.g., *IKZF1, GATA3, NFKB1, RASGRP1, TLR10*), and chromatin organization and cell-cycle control (e.g., *BRD7, E2F4, RMI2*). Additional loci implicated regulators of RNA processing and post-transcriptional control (e.g., *DDX6, ZFP36L1, ATXN2*), consistent with coordinated genetic influences on hematopoietic turnover and stress responses (**Supplementary Table 9**) consistent with coordinated control of hematopoietic function. Notably, the direction of genetic effects revealed marked asymmetry, where majority of shared loci (25 of 47, ∼53%; **Supplementary Data 2**) exhibited opposing effects, such that alleles increasing T1D risk were typically associated with shorter LTL, and vice versa.

**Figure 4.**
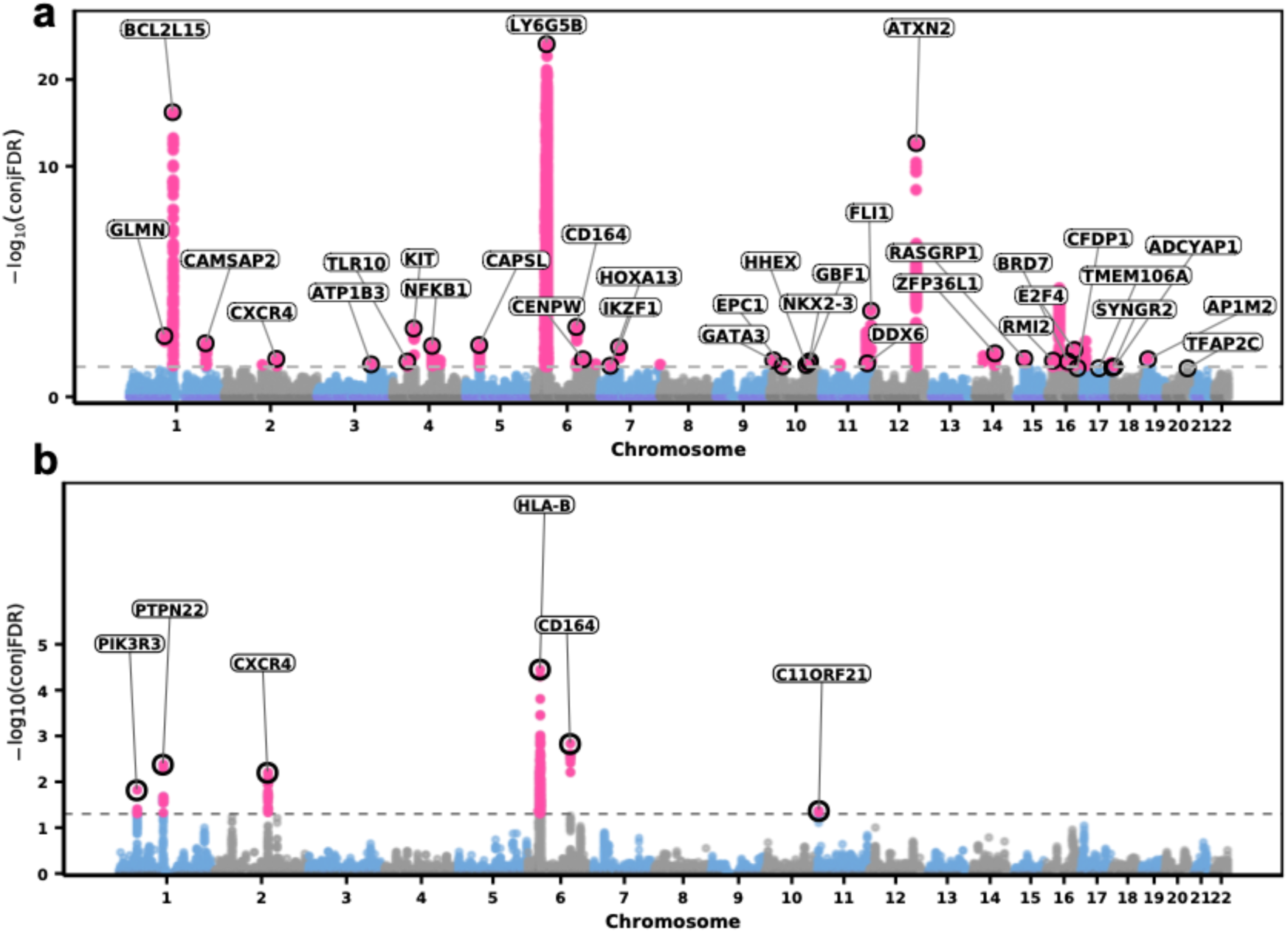
Pervasive genetic relationships between T1D and LTL. (**a**) Genome-wide conjunctional false discovery rate (conjFDR) analysis identifying shared genetic loci between T1D and LTL. Each point represents a genetic variant plotted by chromosomal position and −log₁₀(conjFDR). A total of 47 LD-independent loci were identified at conjFDR < 0.05; to improve visualization, only a subset of lead loci are annotated in the plot. (**b**) ConjFDR analysis of shared genetic loci between T1D and CHIP, demonstrating substantially fewer shared signals compared with LTL.

In contrast to LTL, T1D and CHIP shared few loci outside the MHC region (**Figure 4b**; **Supplementary Data 2**). However, these loci similarly mapped to genes involved in hematopoietic and immune regulation (**Supplementary Table 10**).

To further demonstrate direct involvement of the T1D–LTL loci in HSPC biology, integration with single-cell chromatin accessibility profiles using SCAVENGE revealed a clear enrichment (high trait relevance scores; TRS) in granulocyte–monocyte progenitors (GMPs) (**Figure 5a; Supplementary Data 3**). The highest TRS was in GMPs, followed by common lymphoid progenitors (CLPs), and lower scores in hematopoietic stem cells (HSCs), CMP/LMPP, and erythroid populations.

**Figure 5.**
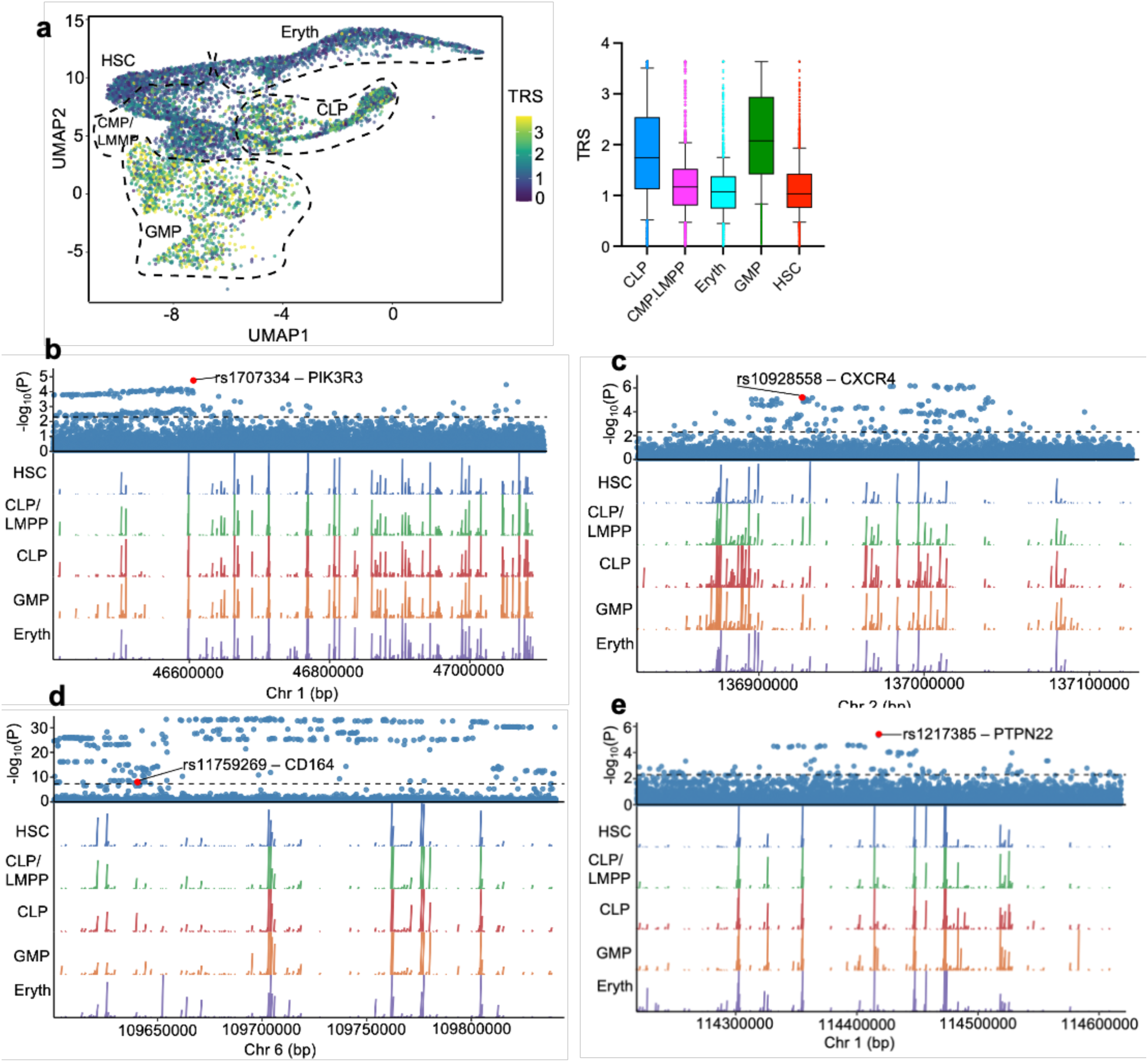
Shared genetic signals between T1D and hematopoietic aging traits preferentially localize to hematopoietic progenitor populations. **(a)** SCAVENGE analysis integrating shared conjFDR loci with single-cell chromatin accessibility profiles from human bone marrow hematopoietic stem and progenitor cells (HSPCs). Left, uniform manifold approximation and projection (UMAP) showing per-cell trait relevance scores (TRS) projected across annotated HSPC populations, including hematopoietic stem cells (HSCs), multipotent/common lymphoid progenitors (CMP/LMPP), common lymphoid progenitors (CLPs), granulocyte–monocyte progenitors (GMPs), and erythroid progenitors (Eryth). Warmer colors indicate higher TRS, reflecting stronger enrichment of shared T1D–LTL genetic signals. Right, distribution of TRS across annotated HSPC populations, demonstrating preferential enrichment in myeloid progenitor populations, particularly GMPs. **(b–e)** Representative shared loci illustrating integration of conjFDR association signals with chromatin accessibility across HSPC populations. Upper panels show local association signals with lead shared variants highlighted in red; dashed horizontal lines denote significance thresholds. Lower panels display aggregated chromatin accessibility tracks across major HSPC populations, showing cell type–specific accessibility surrounding representative shared variants at **PIK3R3** (*b*), **CXCR4** (*c*), **CD164** (*d*), and **PTPN22** (*e*). These loci illustrate convergence of shared genetic associations within regulatory regions active in progenitor compartments implicated by SCAVENGE. Additional nearby variants shown in locus plots represent surrounding GWAS signals for visualization and were not included in SCAVENGE enrichment calculations.

Locus-level integration of four prioritized representative conjFDR loci with chromatin accessibility profiles across HSPC populations further resolved the regulatory architecture of shared genetic signals (**Figure 5b–e**). Three loci (CXCR4, PIK3R3, and CD164) were selected because they were shared across T1D, LTL, and CHIP, representing core pleiotropic signals central to the study hypothesis, whereas PTPN22 was included because of its well-established biological role in T1D. At the CXCR4 locus, the lead shared variant overlapped accessible chromatin with preferential activity in GMPs, consistent with lineage-biased regulation. In contrast, the PIK3R3 locus showed broadly distributed accessibility across progenitor populations, suggesting roles in core hematopoietic regulatory programs. The CD164 locus localized proximal to, but not directly within, accessible chromatin peaks across multiple progenitor states, consistent with distal enhancer-mediated regulation. The PTPN22 locus exhibited prominent accessibility in lymphoid progenitors, in line with its established role in immune regulation. These findings demonstrate that shared loci act through cis-regulatory elements active across hematopoietic progenitors, with a predominant signal in GMPs and additional lineage-specific and distal regulatory patterns.

Last but not the least, Mendelian randomization supported a bidirectional relationship between T1D and LTL (**Figure 6a–b**). Genetic liability to T1D was associated with shorter LTL (IVW: β = −0.0076, 95% CI −0.0125 to −0.0028; p = 0.0019; **Figure 6a**), with consistent directionality across sensitivity analyses. In the reverse direction, genetically predicted longer LTL was associated with reduced T1D risk (IVW: OR 0.83, 95% CI 0.74–0.94; p = 0.0031; **Figure 6b**), although sensitivity estimates were less precise. In contrast, no evidence of a causal relationship was observed between T1D and CHIP in either direction (all p > 0.05). All exposure-associated instrumental variables had F-statistics >10 (range 29.39–2032.35; **Supplementary Data 4**), indicating robust instrument strength with little evidence for weak instrument bias.

**Figure 6.**
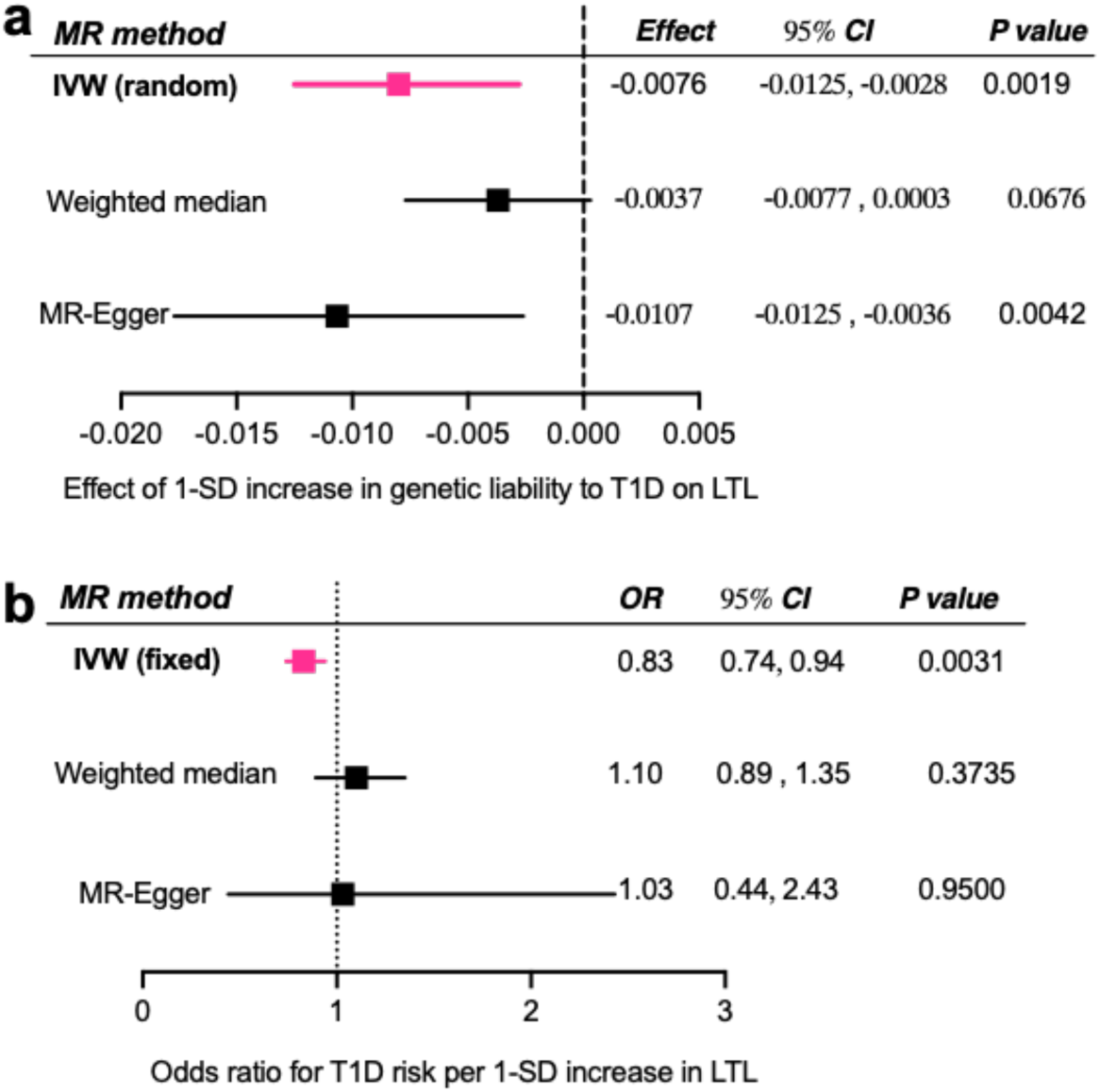
Mendelian randomization (MR) reveals bidirectional relationships between T1D and LTL. (**a)** MR estimates of the effect of genetic liability to T1D on LTL using inverse-variance weighted (IVW), weighted median, and MR-Egger methods. Effect estimates (β, SD units) with 95% confidence intervals (CIs) are shown. (**b**) MR estimates of the effect of genetically predicted LTL on T1D risk using IVW, weighted median, and MR-Egger methods. Odds ratios (ORs) with 95% CIs are shown.

Collectively, these findings support a model in which telomere attrition represents a genetically coupled component of hematopoietic aging in T1D, whereas the increased prevalence of CHIP likely reflects downstream disease-related or environmental processes rather than shared inherited susceptibility.

## Discussion

Hematopoietic aging characterized by clonal hematopoiesis of indeterminate potential (CHIP) and leukocyte telomere length (LTL) attrition has been linked to risk of age-related diseases, including autoimmune and cardiometabolic diseases [25–27,35,36,61,62]. However, these diseases rarely arise abruptly at clinical diagnosis but instead evolve over lifelong genetic predisposition, subclinical immune activation, inflammation, and metabolic stress that shape HSPC turnover, accumulation of somatic mutations and clonal evolution, and telomere attrition [31,63–65]. Therefore, CHIP or shortened LTL observed prior to diagnosis of such conditions may not represent a primary disease-initiating mechanism. In this context, T1D, often diagnosed in children and young adults [37], provides a unique human model to examine how lifelong exposure to immune and metabolic stress reshapes hematopoietic biology and contributes to premature cardiovascular disease.

In this study, we show that T1D is associated with accelerated hematopoietic aging along two biologically distinct axes: higher prevalence and earlier-onset CHIP and disease-accelerated LTL attrition. However, these processes diverge markedly in their relationship to cardiovascular risk. While CHIP is more prevalent and occurs earlier in individuals with T1D, it was not associated with MI or CAD risk or exhibit evidence of meaningful genetic sharing or causal relationship with T1D. In contrast, LTL attrition emerges as a consistent and biologically coupled pathway linking T1D to cardiovascular risk.

In the general population, CHIP has been associated with incident cardiovascular disease, with proposed mechanisms including inflammatory processes that impair vascular and myocardial function [61,65,66]. However, our findings suggest that the contribution of CHIP to cardiovascular risk may be context-dependent. Our genome-wide analyses demonstrated no evidence of a significant shared genetic basis between CHIP and T1D, with genetic correlation estimates that were directionally negative. Emerging evidence suggests that the biological consequences of CHIP may vary across disease contexts. For instance, CHIP has been associated with reduced risk of Alzheimer’s disease [67], highlighting that CHIP may confer protective or compensatory effects in certain settings. These observations suggest that the increased prevalence and earlier-onset CHIP in T1D likely reflect cumulative disease-related perturbations, such as chronic inflammation and metabolic stress, rather than a major driver of cardiovascular risk. Although limited power within the T1D subgroup may have reduced sensitivity to detect modest cardiovascular effects of CHIP, the consistency of largely null epidemiologic and genetic findings argues against a major causal role for CHIP in mediating cardiovascular risk in this setting.

In contrast with CHIP, the relationship between T1D and telomere biology is both stronger and more coherent. While shorter LTL has been associated with cardiovascular disease in the general population [27,62], our findings extend this paradigm by demonstrating that T1D modifies the rate of telomere attrition. Specifically, T1D disease duration is associated with accelerated telomere shortening, indicating that cumulative exposure to immune activation, metabolic stress, and increased hematopoietic turnover drives a disease-accelerated aging trajectory. Genetic analyses further reinforce this connection, revealing extensive overlap and a bidirectional causal relationship between T1D and LTL, including a predominance of opposing allelic effects consistent with antagonistic pleiotropy. This directional architecture suggests that genetic susceptibility to T1D is intrinsically linked to mechanisms promoting hematopoietic turnover and telomere erosion.

The clinical relevance of these findings is underscored by the robust association between shorter LTL and MI within T1D. Our results provide a mechanistic framework for the well-recognized early and excess burden of MI in T1D, raising the possibility that LTL attrition may partially mediate this risk. The weaker and attenuated associations with CAD suggest that telomere biology may be more closely related to processes underlying acute ischemic events than to overall atherosclerotic burden. Preclinical studies have shown that shortened telomeres can induce a shift in hematopoiesis toward myeloid and megakaryocytic lineages, causing imbalances in mature blood cell types [68], processes that heighten inflammation and impair vascular and myocardial function. Furthermore, because LTL correlates with telomere length across multiple somatic tissues, including cardiovascular tissues [69], shorter LTL may also capture telomere-dependent dysfunction within vascular or myocardial cells that could directly contribute to MI susceptibility.

In the general population, CHIP and LTL are often interconnected features of hematopoietic aging [25–27]. Consistent with prior work, CHIP was associated with shorter LTL in the overall cohort. However, this relationship was less evident within T1D. Although statistical interaction was not detected, this apparent dissociation raises the possibility that telomere attrition in T1D is driven predominantly by sustained disease-related stress rather than primarily by clonal expansion. Together with the duration-dependent effects observed for LTL, these findings support a model in which chronic immune and metabolic stress accelerates hematopoietic turnover and telomere erosion largely independently of detectable clonal hematopoiesis.

This study has several strengths, including the use of a large population-based cohort with harmonized CHIP detection and standardized telomere measurements, as well as the integration of epidemiologic, clinical, and genetic analyses. However, several limitations should be considered. The observational design precludes definitive causal inference, and the modest number of individuals with T1D, particularly those harboring CHIP, limits power for gene-specific CHIP analyses and some cardiovascular outcomes. CHIP was assessed at a single time point, precluding evaluation of clonal dynamics over time. In addition, although sensitivity analyses were performed, residual confounding cannot be excluded.

In summary, our findings demonstrate that T1D is associated with accelerated hematopoietic aging but that the biological pathways linking hematopoietic aging to cardiovascular risk are not uniform. Telomere attrition represents a genetically coupled and clinically relevant pathway linking T1D **to** myocardial infarction, whereas clonal hematopoiesis appears to be a secondary, disease-associated process without clear evidence of a direct impact on cardiovascular outcomes in this context. More broadly, these results suggest that the contribution of somatic aging processes to cardiovascular disease is shaped by disease-specific biological contexts, with important implications for risk stratification and mechanistic understanding of vascular disease in T1D.

## Data Availability

The data analyzed in this study were obtained from the UK Biobank under approved application 214427. UK Biobank data are available to qualified researchers upon application to the UK Biobank (https://www.ukbiobank.ac.uk/
). Summary statistics and additional information generated during this study are available from the corresponding author upon reasonable request.

## Supplemental Material

Supplemental Material Tables 1 – 10

Supplemental Material Data 1 – 4

